# Datasets for a Simulated Family-Based Exome-Sequencing Study

**DOI:** 10.1101/2022.05.06.22273576

**Authors:** Nirodha Epasinghege Dona, Jinko Graham

## Abstract

We present simulated exome-sequencing data for 150 families from a North American admixed population, ascertained to contain at least four members affected with lymphoid cancer. These data include information on the ascertained families as well as single-nucleotide variants on the exome of affected family members. We provide a brief overview of the simulation steps and links to the associated software scripts. The resulting data are useful to identify genomic patterns and disease inheritance in families with multiple disease-affected members.

**Specifications Table:** 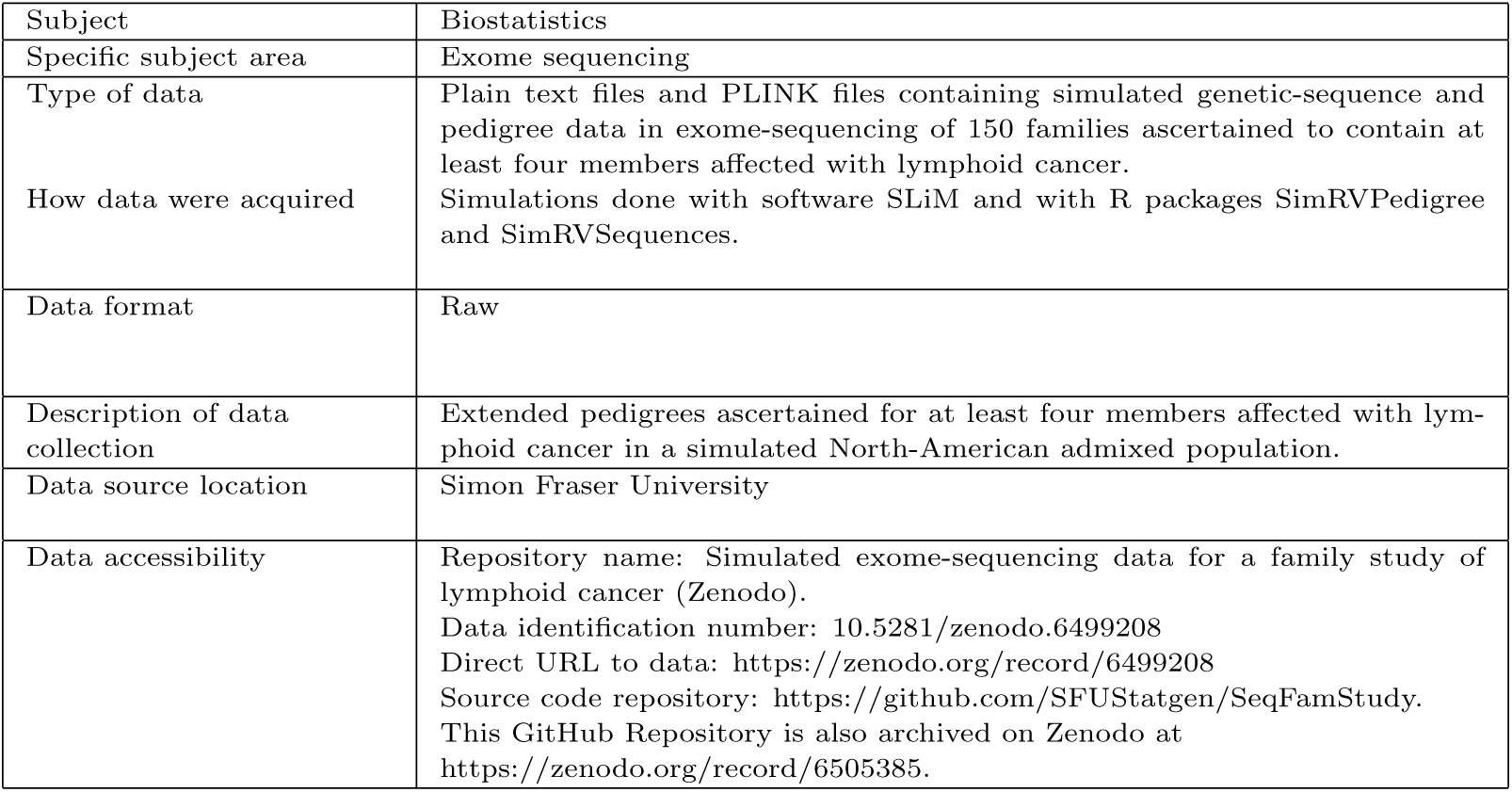

## Value of the Data

- Next-generation sequencing data from families ascertained to contain multiple relatives diagnosed with the same disease can identify rare, causal DNA variants. For example, whole-exome sequencing data has been used to prioritize several rare variants for further investigation in large multi-generational pedigrees ascertained for multiple cases of bipolar disorder [1].
- Data from realistic simulations has the potential to advance the understanding of genomic patterns of disease inheritance, while avoiding costly recruitment of ascertained families and issues of patient confidentiality. Simulated data are easily shared, enabling researchers who analyse sequences collected from ascertained families to test and evaluate different methods.
- We present simulated exome-sequencing data in 150 families ascertained to contain four or more relatives affected with lymphoid cancer. These data should be useful for evaluating genomic patterns and disease inheritance in ascertained families, and for validation and bench-marking of statistical analysis methods in family-based sequencing.
- Our data and simulation scripts answer important calls in genetic epidemiology [2] for the reuse of existing datasets to compare statistical methods and maximize benefit from research investment and for the sharing of source code and simulated data sets in public repositories to facilitate reuse and reproducibility.

## 1. Data Description

This article describes data simulated according to the work-flow in Figure 1 and available in the following files

**Figure 1:**
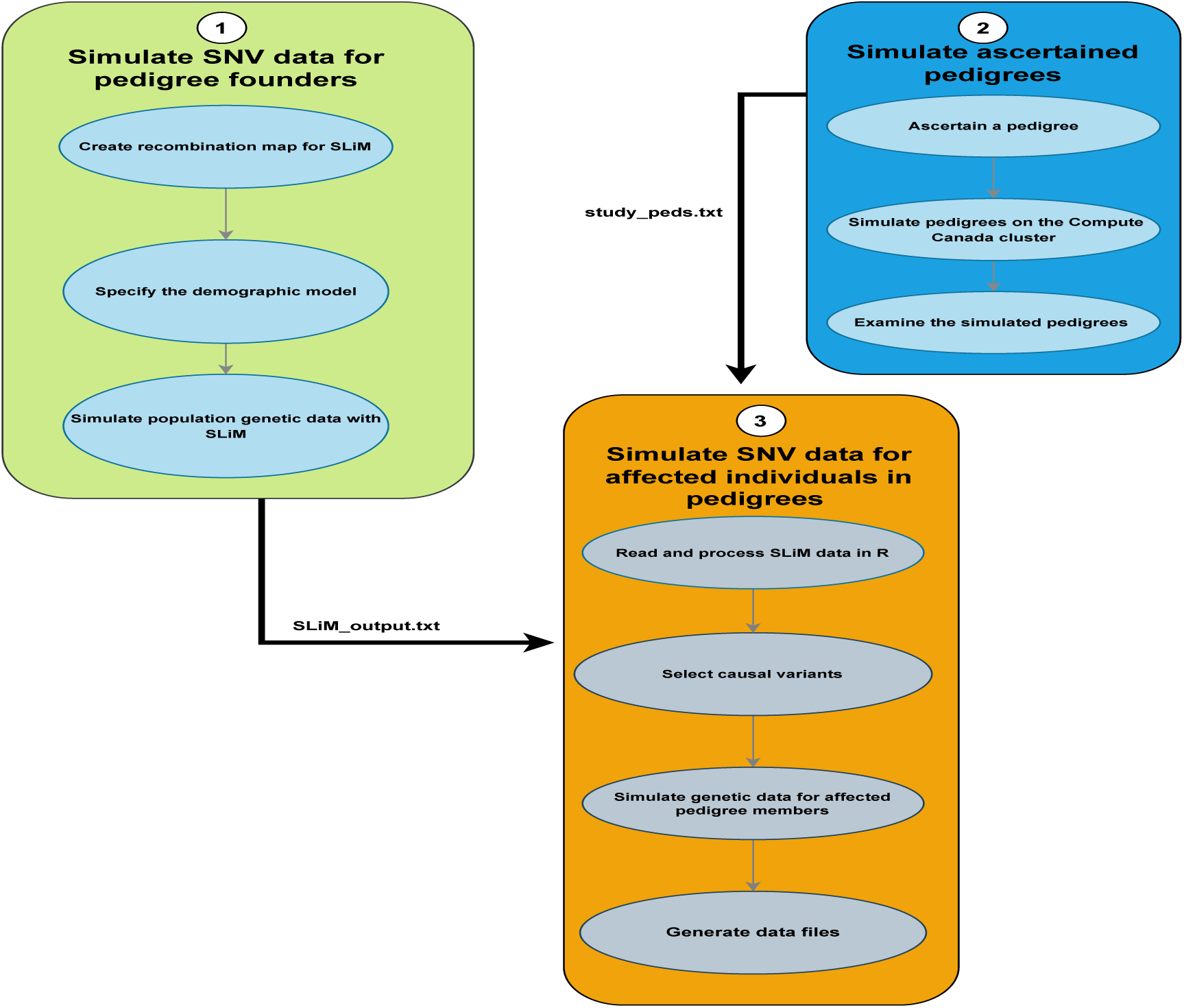
Work-flow for simulating the exome-sequencing data for ascertained pedigrees.

- SLiM output.txt - a 6.3GB text file that contains exome-wide, single-nucleotide variant (SNV) sequences for an American-admixed population of 53876 individuals generated under an American-admixture demographic model [3] with the genetic simulation software SLiM [4].
- SLiM output chr8&9.txt - a 1.3GB text file that contains the SLiM-simulated data above for all source populations as well as the American-admixed sub-population, but only for chromosomes 8 and 9. The total number of individuals in each source population is displayed in Table 2.

**Table 2:** Source Population sizes.
- sample info.txt - a 37.8KB text file giving pedigree information for the disease-affected individuals and individuals connecting them along a line of descent, for all 150 pedigrees. The file contains a total of 1247 individuals, 686 of whom are disease-affected. The remaining 561 individuals in the file connect affected individuals within a pedigree along a line of descent.
- Genotypes.zip - a 5.4MB zip-file that contains 22 chromosome-specific text files of genotypes for rare variants (RVs) on the exome. RVs are defined to be SNVs with population minor-allele frequencies less than 0.01. The RV genotypes are reported in gene-dosage format, as 0, 1 or 2 copies. These files are summarized in Table 3 below.

| Population | size |
| --- | --- |
| African | 14475 |
| European | 35815 |
| Asian | 48765 |
| Admix | 53876 |
| Total | 152931 |

**Table 3:** Number of rare variants (RVs) in each genotype text file.
- SNVmaps.zip - a 2.5MB zip-file that contains 22 chromosome-specific text files giving SNV information. The contents of this zip-file is summarized in Table 4 below.

| Chromosome | 1 | 2 | 3 | 4 | 5 | 6 | 7 | 8 |
| --- | --- | --- | --- | --- | --- | --- | --- | --- |
| No. of RVs | 19393 | 14132 | 11634 | 8096 | 9681 | 10137 | 9653 | 7107 |
| Chromosome | 9 | 10 | 11 | 12 | 13 | 14 | 15 | 16 |
| No. of RVs | 8061 | 8356 | 10629 | 10150 | 4266 | 6108 | 6977 | 7536 |
| Chromosome | 17 | 18 | 19 | 20 | 21 | 22 |  |  |
| No. of RVs | 10168 | 3474 | 10298 | 5007 | 2185 | 4305 |  |  |

**Table 4:** Number of rare variants in apoptosis sub-pathway and number of causal rare variants (cRVs) in SNV map files.
- familial cRV.txt - a 2.4KB text file that contains information about the causal RVs (cRVs) in all 150 ascertained pedigrees. Three of the 150 pedigrees are “sporadic”; i.e., all affected individuals have sporadically occurring disease.
- study peds.txt - a 552.7KB text file that contains the 150 pedigrees ascertained to contain four or more relatives affected with lymphoid cancer.
- PLINKfiles.zip - a 2.8MB zip-file that contains PLINK .fam, .bim and .bed files for all 22 of the chromosomes.
- Chromwide.Rdata - a 217MB .Rdata file that can be used as an intermediate file to save the user substantial time when running the associated RMarkdown script for the simulation. We recommend loading Chromwide.Rdata into your R work-space rather than generating it from scratch.

| Chromosome | 1 | 2 | 3 | 4 | 5 | 6 | 7 | 8 | 9 | 10 | 11 | 12 | 13 |
| --- | --- | --- | --- | --- | --- | --- | --- | --- | --- | --- | --- | --- | --- |
| Pathway RVs | 4 | 90 | 12 | 15 | 24 | 38 | 23 | 50 | 0 | 28 | 5 | 0 | 0 |
| No. of cRVs | 1 | 18 | 4 | 3 | 7 | 7 | 5 | 8 | 0 | 6 | 2 | 0 | 0 |
| Chromosome | 14 | 15 | 16 | 17 | 18 | 19 | 20 | 21 | 22 |  |  |  |  |
| Pathway RVs | 0 | 0 | 8 | 19 | 30 | 6 | 0 | 37 | 18 |  |  |  |  |
| No. of cRVs | 0 | 0 | 3 | 3 | 5 | 0 | 0 | 3 | 5 |  |  |  |  |

Figure 2 shows an example family (family ID 39 out of 150) from study peds.txt. This family contains 16 individuals across 4 generations, 4 of whom are affected (IDs 1, 4, 8 and 10). Across all 150 families, the family size varies between 8 and 207 individuals (see Figure 3), the number of affected members varies between 4 and 8 individuals (see Figure 4) and the number of generations varies between 1 and 7 (see Figure 6). Further, as the number of generations increases, so does the pedigree size (see Figure 7). Note that, due to high computational cost, we did these calculations only for a single set of 150 pedigrees. However, we can increase the number of replications in this simulation and a detailed description about how users can generate more pedigrees can be found in our second supplementary material.

**Figure 2:**
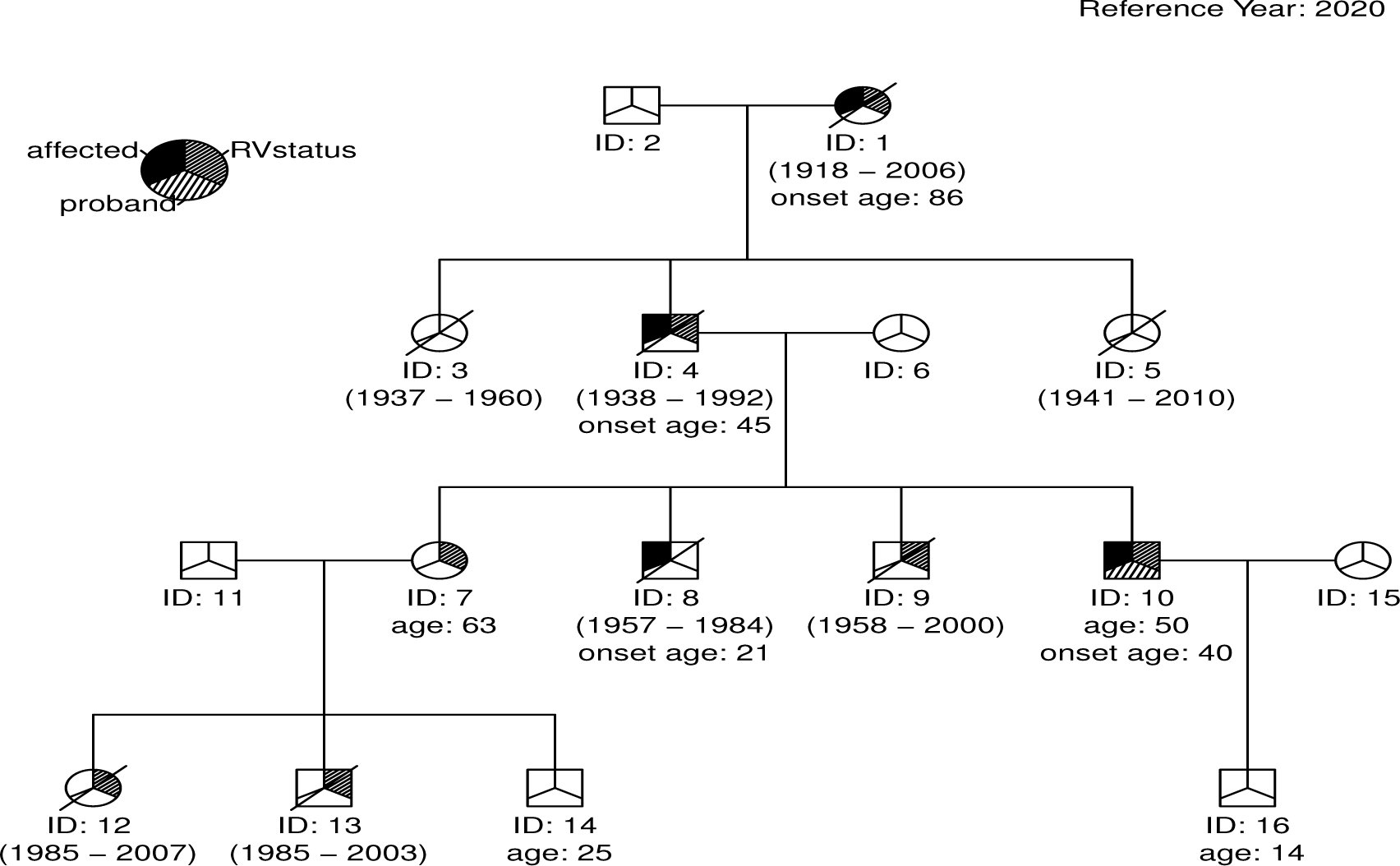
An example pedigree (pedigree 39 out of 150). The legend identifies affected individuals, the proband, and the cRV status of the individuals. Disease-affected individuals have solid shading in the upper-left third of their symbol (IDs 1, 4, 8 and 10). The proband (ID 10) has shading in the lower portion of their symbol. Individuals carrying a causal genetic variant (IDs 1, 4, 7, 9, 10, 12 and 13) have shading in the upper-right portion of their symbol. The birth year and the death year of dead individuals are displayed in parentheses. The age of the individuals who are alive at the end of the reference year of 2020 displays under their symbol. Any individual with disease onset before the end of the reference year has a disease-onset year given under their symbol. Following standard practice in medical genetics, individuals who have died as of the reference year have slashes through their symbols. The age of the individuals who are alive at the end of the reference year displays under their symbol. Any individual with disease onset before the end of the reference year has a disease-onset year given under their symbol.

**Figure 3:**
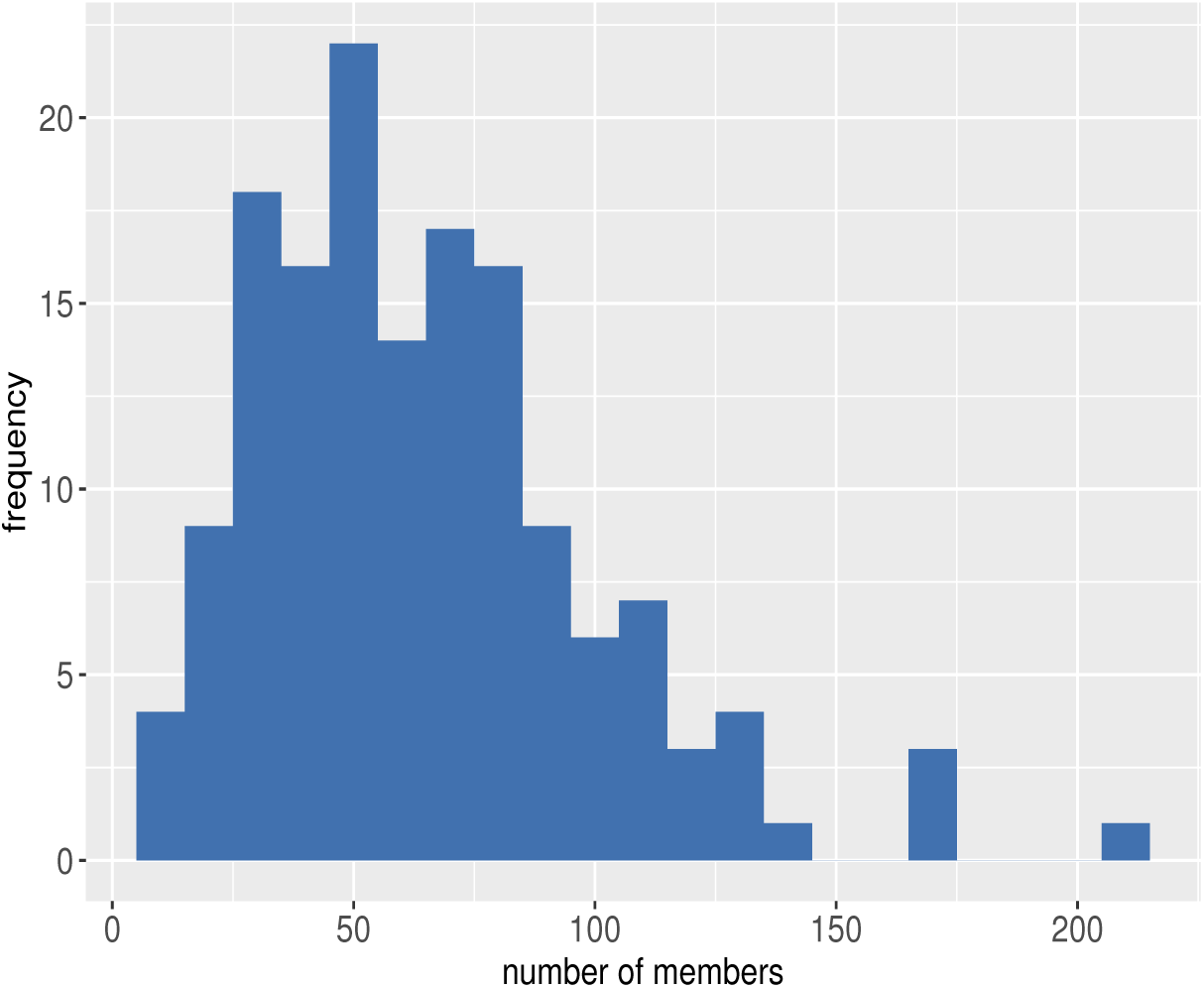
Distribution of pedigree size.

**Figure 4:**
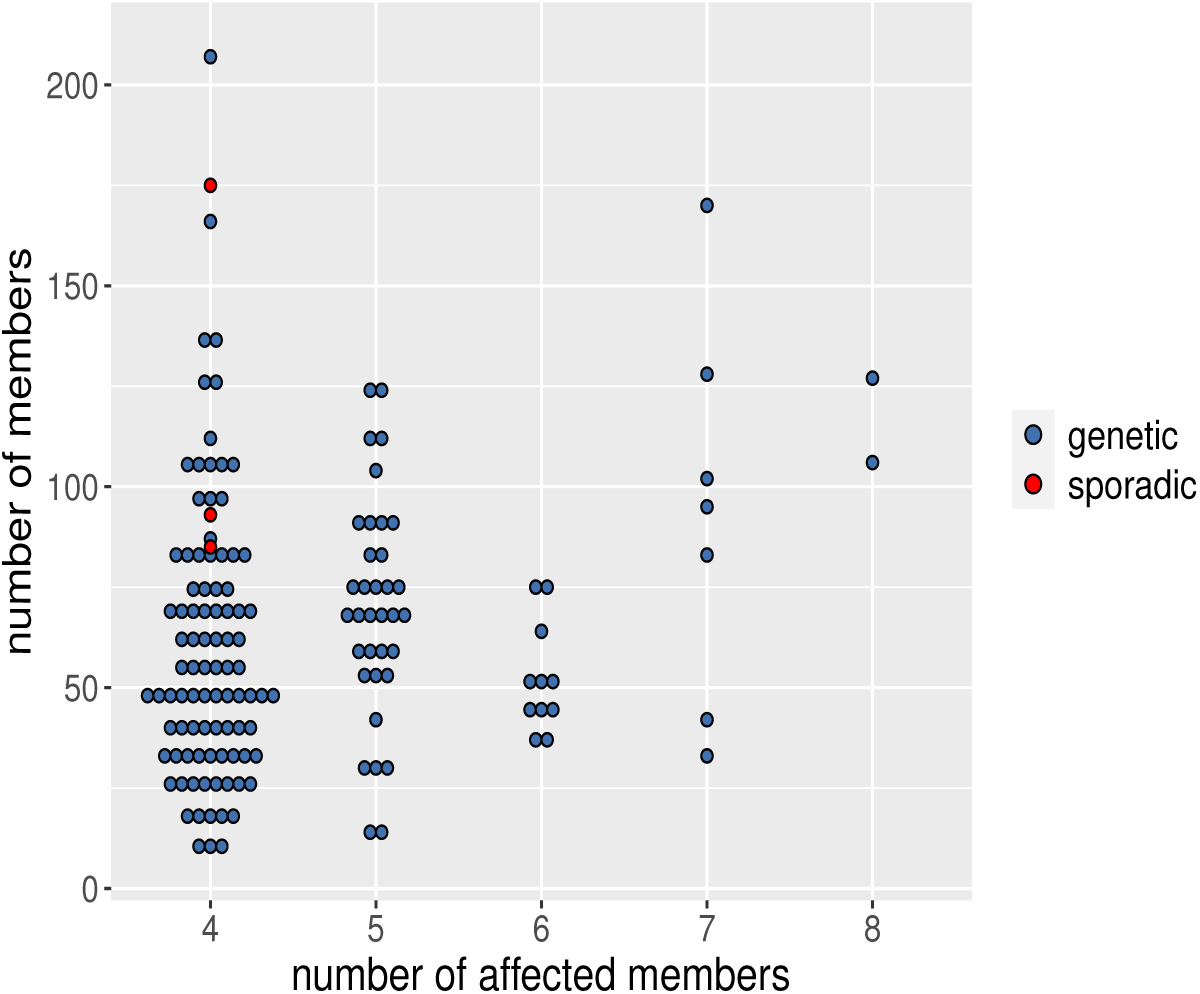
Distribution of pedigree size by number of affected members.

**Figure 5:**
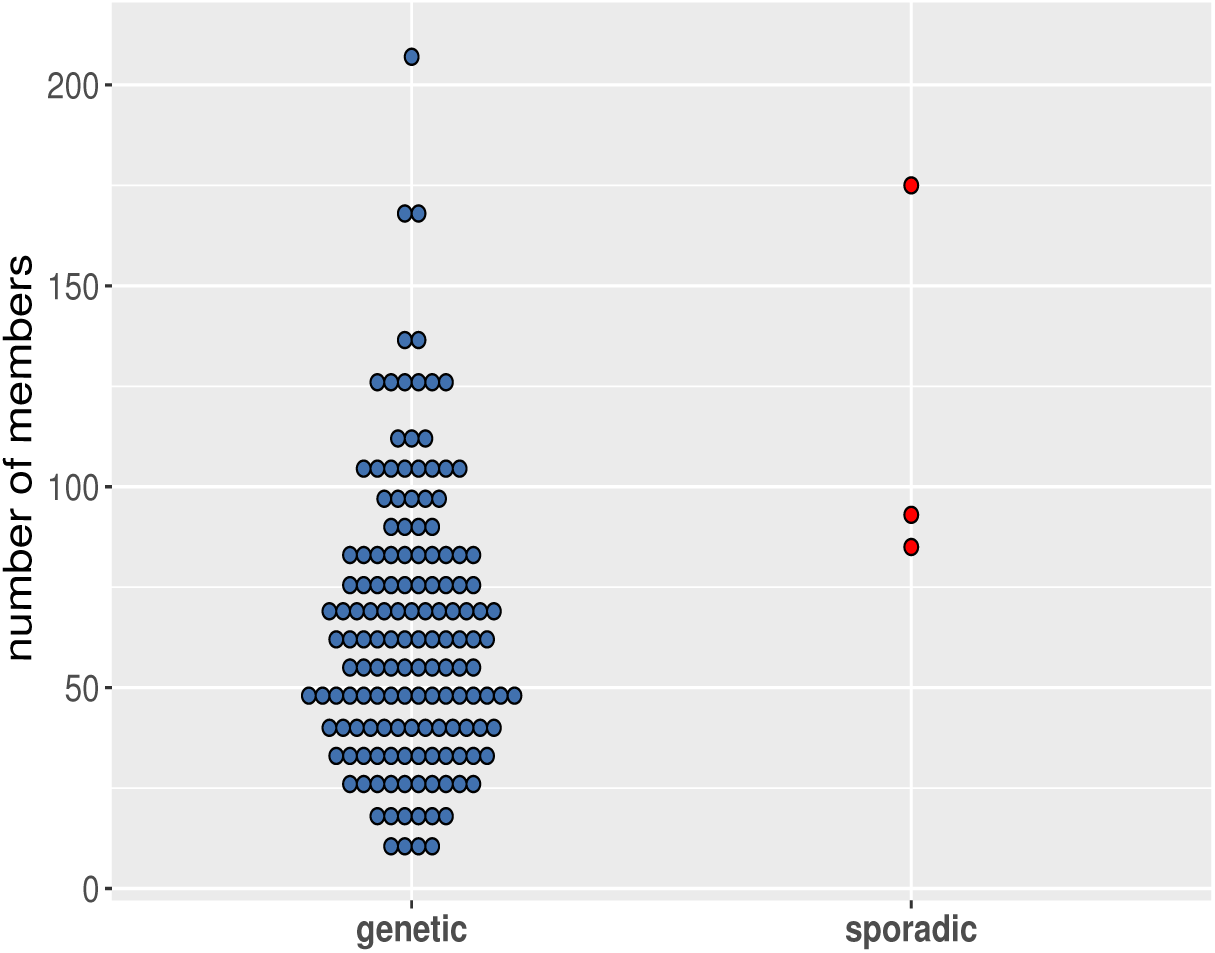
Distribution of pedigree size in pedigrees with genetic and sporadically occurring disease.

**Figure 6:**
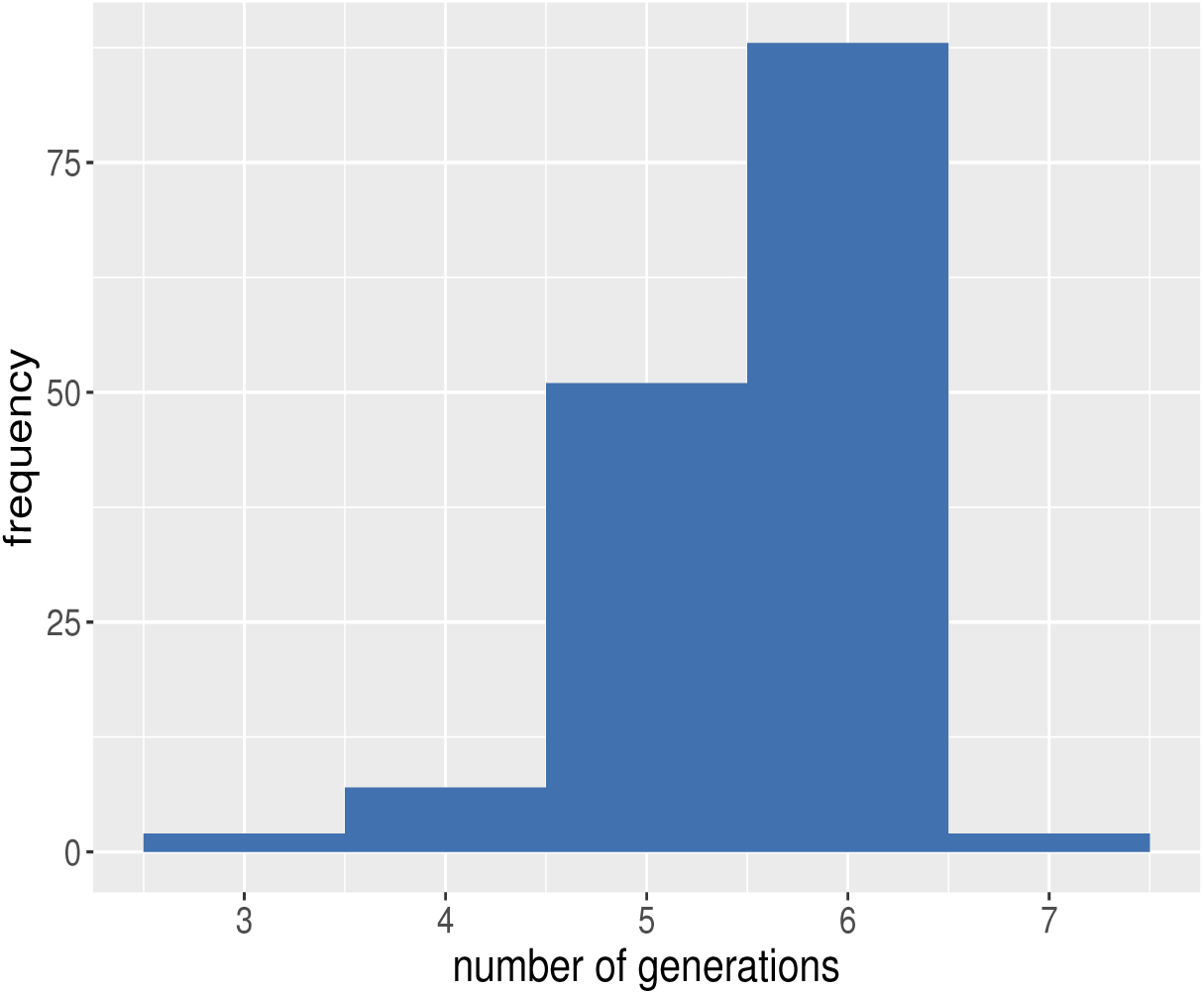
Distribution of number of generations in pedigrees.

**Figure 7:**
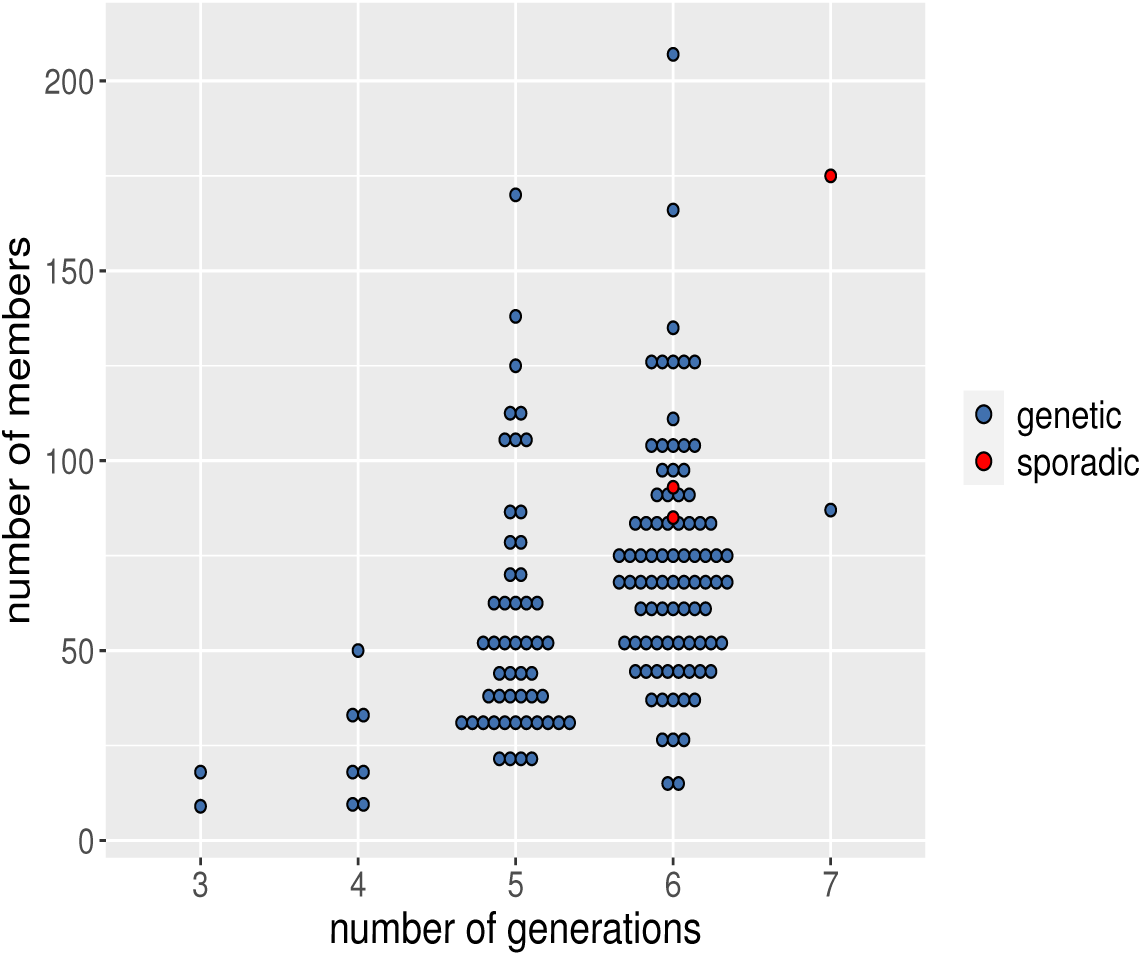
Distribution of pedigree size by the number of generations.

Figure 4 suggests that the size of the pedigrees is uncorrelated with the number of affected members. Also, as expected and shown in Figure 5, so-called “sporadic” pedigrees, in which all affected members have sporadically occurring disease (family IDs 72, 95, 103), tend to be larger than non-sporadic pedigrees.

The above data files have been created by simulation, performed in three major steps as outlined in Figure 1 and implemented in the RMarkdown files provided as supplementary materials. The RMarkdown files to simulate the data are

- Supplementary Material 1-A: Simulate SNV sequence data for pedigree founders. Contains the code and methods to get the `SLiM_output.txt` file.
- Supplementary Material 1-B: Combining Source Populations with the American-Admixed Population in SLiM. Contains the code and methods to get the `SLiM_output chr8&9.txt` file.
- Supplementary Material 2: Simulate ascertained pedigrees. Contains the code and methods to get the `study_peds.txt` file.
- Supplementary Material 3:Simulate SNV data for affected individuals in pedigrees. Contains the code and methods to get the data files `sample_info.txt, Genotypes.zip, SNVmaps.zip familial_cRV.txt` and `PLINKfiles.zip`. Also contains a detailed description of the formats of these files.

## 2. Experimental Design, Materials and Methods

To acquire these data, we simulated exome sequences in pedigrees ascertained to have four or more relatives affected with lymphoid cancer, according to the work-flow summarized in Figure 1. The simulation involved three major steps which we describe in the subsections below.

### 2.1. Simulate SNV data for pedigree founders

Simulating the exome sequences in ascertained pedigrees requires SNV sequences for pedigree founders. We assume these founders are sampled randomly from an American-Admixed population, which we simulate with the evolutionary simulation package SLiM [4]. To mimic exome sequencing, we simulate genome-wide sequences of exons only. A detailed account of this part of the simulation is provided in Supplementary Material 1-A.

As shown in Figure 1, simulating the founder sequences involves three steps. First, we provide a recombination map to SLiM giving the exon positions in chromosomes. We use the `create_SlimMap()` function in the `SimRVSequence` [5] R package for this task.

Second, we specify a demographic model for the American-Admixed population [3] because our work is motivated by a family-based exome-sequencing study of lymphoid cancer in a North American population [6]. The American-Admixture demographic model of [3] is compiled in `stdpopsim`, a standard library of population-genetic simulation models [7].

Finally, we use the Compute Canada cluster (http://www.computecanada.ca) to avoid the computational cost of running this large simulation on a personal computer. The batch scripts that we ran on the cluster can be found in the first supplementary materials. The resulting SNV data for pedigree founders is used as an input to simulate the SNV data for affected pedigree members in the third step of our work flow, described below.

### 2.2. Simulate ascertained pedigrees

The second step of our workflow is to simulate ascertained pedigrees with disease-affected relatives. We simulated 150 pedigrees ascertained to contain four or more relatives affected with lymphoid cancer using the `SimRVPedigree` R package [8]. A complete description of this step of the workflow can be found in our Supplementary Material 2. As the simulation is time-consuming, we use the Compute Canada cluster. The final outcome of this step is a set of ascertained pedigrees in which to generate exome-sequencing data for the third and final step of the workflow, described next.

### 2.3. Simulate SNV data for affected individuals in pedigrees

To simulate exome-sequencing data for the affected members of ascertained pedigrees, we use the outcomes from the previous two steps of the workflow, together with the gene-dropping functions in the `SimRVSequences` R package [5]. These functions require sparse matrices of SNV sequences but, unfortunately, the population size and number of SNVs is too large to accommodate within a single sparse matrix. Therefore, we create the sparse matrices chromosome-by-chromosome. A complete description of how we implemented this final step of our workflow can be found in Supplementary Material 3.

As shown in the Figure 1, we divide this step into four sections. First we read the SLiM-simulated sequencing data into R and process them chromosome-by-chromosome. Then we select the rare variants (RVs) in genes on an apoptosis sub-pathway as candidates for causal rare variants (cRVs), as described in [5]. Specifically, we use the apoptosis sub-pathway centered about the TNFSF10 gene in the UCSC Genome Browser’s Gene Interaction Tool. This sub-pathway contains 23 genes: *DAP3, CFLAR, CASP10, CASP8, TNFSF10, CASP3, MAP3K1, TNF TNFRSF10A, FOXO3, IFNGR1, CYCS, TNFRSF10B, TNFRSF11B, FAS, FADD, TRADD, TP53, RALBP1, BCL2, BAX, IFNAR1, IFNGR2* and *BID*. These genes and their chromosomal locations are provided in the hg apopPath data set of the SimRVSequences package. Among the SNVs in this pathway, cRVs are selected from population singletons, on the basis of their absolute selection coefficients, until the cumulative probability of a sequence carrying any cRV is 0.001.

We then simulate exome sequences for the affected individuals in the 150 ascertained pedigrees by using the `SimRVSequences` R package. The `sim_RVstudy()` function of this package simulates genetic sequence data in pedigrees, but expects only a single population database of sequences as an argument in the form of a sparse matrix of SNV haplotypes and an associated mutation data frame. Unfortunately, we cannot use `sim_RVstudy()` without modification because the number of individuals and RVs in our American-admixed population is too large. The fundamental problem is that the population sequences of RVs cannot be contained in a single sparse matrix without exceeding the memory capacity of R. We therefore modify `sim_RVstudy()` and various supporting functions in the `SimRVSequences` package to handle chromosome-specific databases as described in the Supplementary Material 3.

Finally, we deliver our data in human-readable flat-file formats. We use the simulated data to create a .sam file containing information about genotyped individuals in the ascertained pedigrees, chromosome-specific .geno files containing RV genotypes and chromosome-specific .var files containing information about RVs. The data files are in flat-file format and also in PLINK file formats [9]. These text and PLINK file formats are discussed in the Supplementary Material 3.

## Data Availability

All data produced are available online in the referenced Zenodo repository. Also, the code used to generate the data is available in a GitHub Repository that has been archived on Zenodo as well.

https://zenodo.org/record/6499208

https://zenodo.org/record/6505385

## Acknowledgments

This research was supported by the Natural Sciences and Engineering Research Council of Canada (NSERC), RGPIN/04296-2018 and used the computational resources provided by Compute Canada (http://www.computecanada.ca).

## CRediT Author Statement

**Nirodha Epasinghege Dona**: Conceptualization, Methodology, Software, Validation, Formal analysis, Investigation, Data Curation, Writing-Original Draft, Visualization.

**Jinko Graham**: Conceptualization, Methodology, Resources, Writing-Review & Editing, Supervision, Funding acquisition.

